# Dreams in Blind individuals: A mystery of brain and vision

**DOI:** 10.1101/2023.03.10.23286640

**Authors:** Dev Desai

## Abstract

**Background:** Dreaming and synthesizing a dream world is a complex process undertaken by the brain with the impulses it has received. It’s often said that blind people can’t see in their dreams but some papers suggest that they in fact do see in their dreams. This knowledge can be used to understand how actually the dream world works and get more insights about the whole process.

**Aim:** To identify if different categories of blind individuals can see or not and to understand if they can dream, what type of dreams they get and whether they have vision in their dreams or not.

**Methodology:** A cross sectional survey based study was conducted with interview pattern to identify the patterns of dreaming. Questions about duration since they have visual impairment were also asked and data was digitalized and later analyzed in excel.

**Results:** A total of 75 visually impaired individuals were recruited for the study who were willing to give consent for the study. It was seen that individuals who lost their sight totally before attaining a level of mental maturity that they can comprehend the concept of light were unable to see in their dreams. Individuals who lost their sight completely after attaining that level of maturity could see in their dreams but the vision in their dreams was only limited to what they had seen and what they could imagine from that limited experience of light sense. Other individuals who still could percept light could see in their dreams what they could see in the real word.

**Conclusion:** Visually impaired individuals can in fact see in their dreams provided they still can percept light and see and if they have lost their sight completely then they lost it after gaining enough mental image maturity to understand the concept of light.

## Introduction

Dreams and Dream world is a place about which we know very little and it fascinates humans till these days. Dreaming is an integral part of one’s sleep cycle and for some people it is an source of motivation, for others a source of ideas and for some people a place where they can do anything and they feel like a king in it. (1) Some people are also there who don’t like dreaming at all and it makes them tired and for some people nightmares are the hardest thing they have to endure in the entire day. All in all, dreaming and the experience may be one of the most fascinating thing. (2)

Physiologically, dreaming is a phenomenon which occurs in REM (Random Eye Movements) part of the sleep which occurs on an average almost every 90 minutes of sleep and is 20 minutes long. (3) Dreaming is considered by scientists and Dream analysts as a way for the brain to reorganize the experiences and thoughts of the day and convert the short term memory into long term memory and to relax and remove the toxins it accumulated in the entire day.

While Dreaming, it’s almost like the mind is dissociated from the body but the eyes are not and the eyes move rapidly with eyelids closed, hence it’s called REM sleep. (4)

It’s been observed and documented enough that a person only greets the faces it has seen in real life, only sees the places its already been to, and only experiences the feeling and experiences it has already felt. There is still a debate that whether blind individuals can see in their dreams or not and if not, then what do they dream about, because a dream of normal individuals mostly consists of vision along with sound and touch with less amount of smell and taste. (5)

A hypothesis can be generated here, with the above facts that if a blind person has experienced what light looks likes and if they can contemplate the concept of vision, then they can see in their dreams but then the question arises of till what limit they can see in their dreams, and if they can see then what do they see in it.

The main question is whether totally Blind individuals see anything in their dreams especially those who have been visually impaired since birth, and if they can’t see in their dreams then what does their dream consist of.

The reason behind this study is to once and for all answer the questions about dreams in blind individuals.

A Survey based Cross sectional study was conducted with 75 visually impaired individuals was conducted. Along with demographical details, questions about whether they dreamt or not and if they dreamt, then what does these dreams consist of was asked. Questions about their visual impairment along with for how long they have been visually impaired were asked. Every visually impaired blind individual have been given a certificate about the degree of the disability, which were checked and these individuals for the sake of analysis were divided into different categories.

## Inclusion criteria

- Individuals agreeing to take part in the study and giving their consent to answer the questions
- Individuals having their disability card given to them by the government actually showing the degree of disability they suffer with

The data was collected in a interview based system and then the data was digitalized in excel and proper statistical tools were used for analysis.

## Results

**Figure 1:**
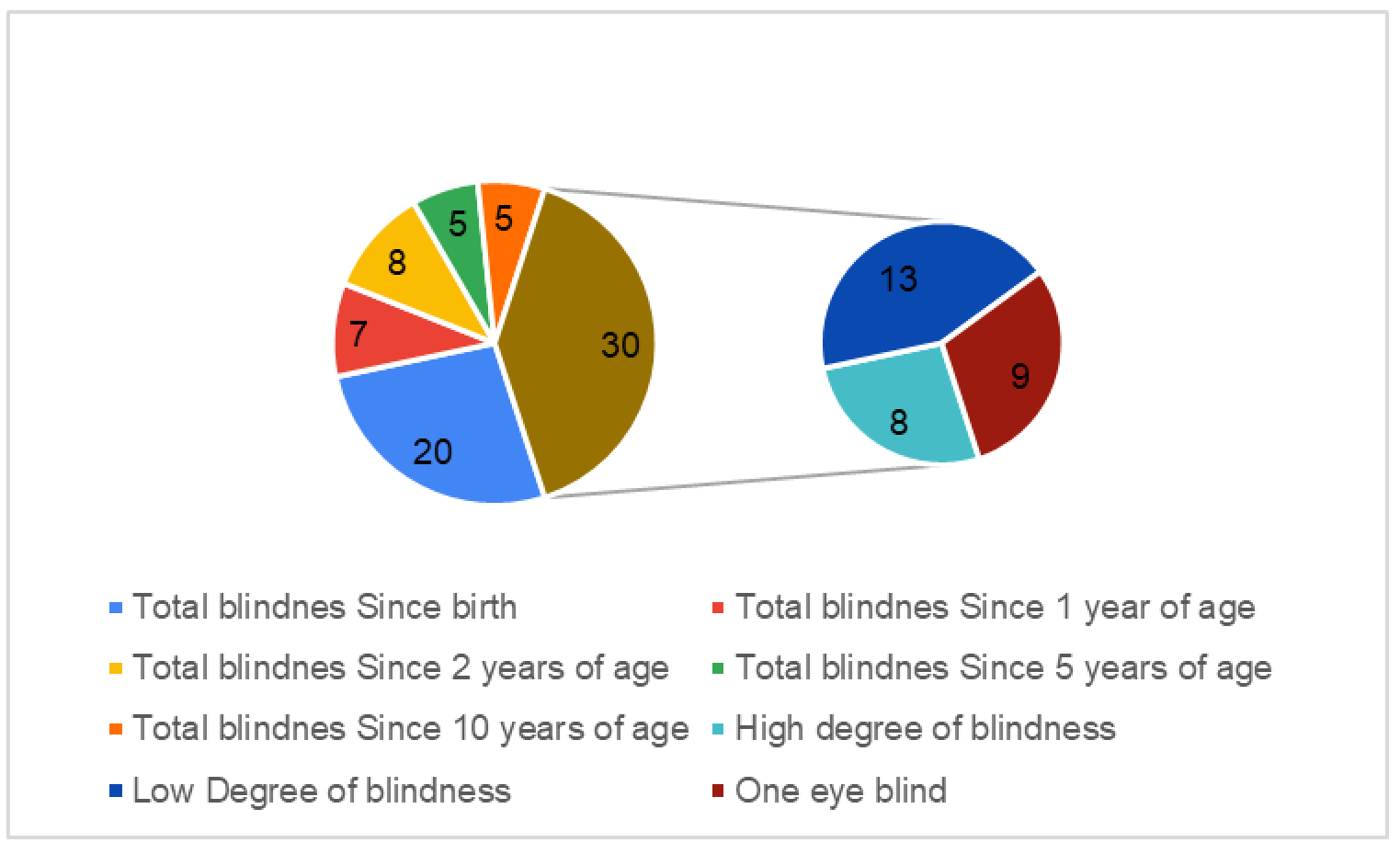
Distribution of Individuals.

**Table 2:** Do they see in their Dream ??

| Table 2:- Do they see in their Dream ?? |  |
| --- | --- |
| Total blindness Since birth | No |
| Total blindness Since 1 year of age | No |
| Total blindness Since 2 years of age | No |
| Total blindness Since 5 years of age | Yes |
| Total blindness Since 10 years of age | Yes |
| High degree of blindness | Yes |
| Low Degree of blindness | Yes |
| One eye blind | Yes |

## Discussion

It can be seen here with the results here that people who have retained even a slight amount of vision and can actually see in their dreams and have vision while they are dreaming. It means that the people who have high degree of visual impairment, Low Degree of visual impairment, and one eye blindness can see. It shows that the people who can perceive light can see in their dreams and have dreams like normal people do. When asked, it was found that people with high degree of visual impairment who have impairment since very low age or from birth and have not registered light as a proper impulse after gaining mental age maturity, and now only have perception of light and perception of direction, they said that they could only see the flashes of light in their dreams as they see when they are awake. People with low degree of blindness agreed that they could only see with the clarity they can see when they are awake. They also agreed that they could see in their dreams but it was very limited. One eye blind individual with the other eye normal or near to normal with external help like spectacles and contact lenses showed that they had dreams where they could see and their dreams were like normal individuals with normal vision.

People who were totally blind could be divided into three parts. The individuals from this group who were blind since birth and have never ever known what light is and what light feels like on their retina, they showed that they in fact could not see in their dreams and their dreams only consisted of hearing, taste, smell and touch, meaning that they were blind in their dreams too. (6) (5) The other group who lost their vision after birth but before gaining mental age maturity that their brain could remember what light is, they also said that they could not see in their dreams. It means that not only totally blind since birth but individuals who were blind from very young age, they also cannot see in their dreams. The third group comprises of individuals who had lost their visions after gaining mental age maturity so that their brain could register what light is and how to process a light impulse if a light particle falls on retina and the whole visual afferent pathway and brain have no dysfunctions. These individuals’ despite being totally blind could actually see in their dreams. The peculiarity of these dreams were that they could see only the things they had seen when they had vision and they could not synthesize or imagine new places to go in their dreams. Most of the individuals from this group said that the most frequent thing that they see in their dreams is their mother’ face. (7) (8) (9) (10)

These results all tell a story. A story of how dreams work. It shows that for the brain to synthesis a dream, first it needs the raw material which are the impulses from the senses however old they may be. The brain cannot recreate its own impulses to put in the dream as the people with total blindness since birth cannot see in their dreams. Secondly, to register the impulse, the brain needs to be mature of that level. Physical age can be different when a child attains this hypothetical mental age maturity or it may be a continuous process which runs for a unknown and variable period of time where child learns to register and remember this impulses from easiest to hardest across all senses simultaneously. Thirdly, brain can use these registered impulses to create dreams as it seems fit. Fourthly, to experience any impulse, for the sake of argument vision, a person needs to know the impulse. For an example, a person can’t go to places he or she have never visited nor imagined nor have seen it. A impulse of that place and an impulse of the existence of the place even though imagined are a must to reproduce that place in the dream. Same goes with faces. If a person has never seen someone’s face and don’t know they exist, then their brain cannot reproduce that face in the dream. So to answer the question and talk about the hypothesis, it seems like the hypothesis may be true that if the person has the knowledge of what light is and what it feels like and can grasp the concept of light, not theoretically but practically first-handedly, then it seems like they can see in their dreams of what they have experienced otherwise without any of the component mentioned above, they will not be able to synthesis a dream with vision in it.

## Conclusion

It can be concluded here that visually impaired individuals can dream provided they understand what is light and can grasp the concept of light. Individuals who don’t know what light is, like totally blind individuals since birth and individuals who lost their sight completely before attaining mental age maturity, cannot in fact see in their dreams and their dreams comprises of mainly sound along with touch and taste. It was also found that not only blind individuals but the same concept applies to normal individuals also when it comes to fundamentals of building a dream world that any impulses which doesn’t exist for an individual will not be synthesized in the dream and that dream can only be made of the impulses experienced or imagined with full consciousness.

## Data Availability

All data produced in the present study are available upon reasonable request to the authors

